# Safety and immunogenicity of Pfizer/BioNTech SARS-CoV-2 mRNA third booster vaccine against SARS-CoV-2 Omicron variant in Japanese healthcare workers

**DOI:** 10.1101/2022.01.20.22269587

**Authors:** Yohei Seki, Yasuo Yoshihara, Kiyoko Nojima, Haruka Momose, Shuetsu Fukushi, Saya Moriyama, Ayumi Wagatsuma, Narumi Numata, Kyohei Sasaki, Tomoyo Kuzuoka, Yoshiyuki Yato, Yoshimasa Takahashi, Ken Maeda, Tadaki Suzuki, Takuo Mizukami, Isao Hamaguchi

## Abstract

**Background:** The Omicron variant of severe acute respiratory syndrome coronavirus 2 (SARS-CoV-2) was identified in Japan in November 2021. This variant contains up to 36 mutations in the spike protein, the target of neutralizing antibodies, and can escape vaccine-induced immunity. The third booster vaccination campaign began with healthcare workers and high-risk groups. The safety and immunogenicity of third booster vaccination against Omicrons remain unknown.

**Methods:** In total, 272 healthcare workers were evaluated for their long-term safety and immunogenicity. Here, we established vaccine panels to evaluate the safety and immunogenicity against variants of concern (VOCs), including the Omicron variant, using a live virus microneutralization assay.

**Findings:** Two-dose vaccination induced robust anti-spike antibodies and neutralization titers (NTs) against the ancestral strain WK-521, whereas NTs in VOCs were significantly decreased. Within 93–247 days of the second vaccine dose, NTs against Omicron were completely abolished in up to 80% of individuals among the vaccine panels. The third booster vaccination induced a robust increase in anti-spike antibodies and NTs against the WK-521, Delta, and Omicron variants. The breadth of humoral immunity and cross-reactivity with Omicron increased. The cytokine signature and adverse event rate remained unchanged after three-dose vaccination.

**Conclusions:** The third vaccination dose is safe and effective against Omicron infection.

**Funding:** This study was supported by grants from AMED (Grant Number JP21fk0108104 and JP21mk0102146).

## Introduction

Severe acute respiratory syndrome coronavirus 2 (SARS-CoV-2) emerged at the end of 2019, in Wuhan, China, and rapidly spread worldwide.^1^ The disease was declared by the World Health Organization (WHO) as a pandemic in March 2020.^2^ In Japan, the first coronavirus 2019 (COVID-19) case was reported in January^3^ and the second in February 2020, after which a large number of COVID-19 cases were reported from the Diamond Princess cruise ship at Yokohama port near Tokyo, Japan.^4^ The number of infections increased slightly within cities but the first major wave of infection of more than 10,000 cases per day occurred in April 2020. The Japanese government declared its first state of emergency in major cities, including Tokyo, and implemented restrictive measures. As of January 19, 2022, the Japanese government has declared a state of emergency four times following the five pandemic waves in Japan.^5^ During the second wave of the pandemic, SARS-CoV-2 acquired the D614G mutation in the spike region, affecting its transmissibility^6^ and infectivity.^7,8^ The newly emerged Alpha strain (B.1.1.7) was isolated from the UK and contained an additional E484K mutation, which gradually appeared to increase and replace the dominant endemic virus.^9^ Simultaneously, the Beta strain was isolated from South Africa with a new K417 mutation, along with three of the mutations observed in the Alpha variant (E484K, N501Y, and D614G in the spike protein), enabling the virus to escape vaccine-induced antibodies.^10^ The Gamma (P.1) variant with a new K417T mutation and three previously identified mutations observed in Alpha was also isolated in Brazil.^11^ The WHO defined variants of concern (VOCs) as those that may impact society. In late summer 2020, many countries began reviewing and authorizing new types of vaccines, such as the Pfizer/BioNTech SARS-COV-2 mRNA vaccine,^12^ Moderna mRNA vaccine,^13^ Janssen/Johnson & Johnson adenovirus vectored DNA vaccine,^14^ and AstraZeneca adenovirus vector DNA vaccine.^15^ In December 2020, the US and EU countries approved these vaccines under Emergency Use Authorization. These SARS-CoV-2 vaccines have remarkably reduced the number of COVID-19 infections, hospitalizations, and deaths in clinical trials in many countries.^16^ The Japanese government first approved two mRNA vaccines in February 2021, after which vaccination of healthcare workers and high-risk people aged over 65 years was initiated. The Murayama Medical Hospital of Medical Hospital Organization is a major Japanese national organization that began two-dose vaccination as part of post-licensure safety and immunogenicity surveillance. The National Institute of Infectious Diseases (NIID) collaborated with Murayama Medical Hospital of Medical Hospital Organization to evaluate neutralization titers (NTs) by performing live virus neutralization assays to compare their anti-spike antibody and adverse event rates. Major surveillance is ongoing; however, a vaccine panel is required to help understand the effects of vaccination against newly emerged VOCs in a limited number of participants.

In Japan, since August 2021, the Delta variant (B.1.617.2), which has an L452R mutation and previously identified D614G mutations, has emerged and become dominant in Japan. Because of the neutralization ability induced by mRNA vaccination,^17^ the number of daily Delta variant infections has gradually decreased in Japan. In November 2021, the SARS-CoV-2 Omicron variant (BA.1/B.1.1.529) first emerged in Botswana and caused a large number of infections in South Africa. The WHO and, soon after, the Japanese government, declared the Omicron variant as a novel VOC. This strain contains more than 36 mutations in the spike protein, for which vaccines appear to be less effective at inducing immunity. A recent study showed that mutations in the receptor-binding domain (RBD) of spike protein enable escape from vaccine-induced immunity^18–20^ and increase infectivity by enhancing the affinity for angiotensin-converting enzymes (Tian et al., 2021). In addition, the Omicron RBD has 15 mutations, some of which overlap with previously reported variant mutations, such as K417, E484, and N501 in Beta (B.1.351) and Gamma (P.1).^19^ A recent study showed that the Omicron spike evasion of the virus to the Pfizer/BioNTech mRNA vaccine is 44-fold more efficient than that of the Delta variant.^21^

Several studies have suggested that antibodies against SARS-CoV-2 gradually decrease after the second vaccination,^22^ even by 6 months after two-dose mRNA vaccination induced durable immune memory to SARS-COV-2 variant of concerns.^24^ Thus, introduction of a third booster vaccination may be beneficial. Some studies suggested that booster immunization is effective against the Omicron variant according to a pseudovirus neutralization assay.^23^ In addition, a heterologous booster can induce greater immune responses and enhance immune protection compared to homologous vaccinations, and^25^ the immune-boosting strategy is safe for healthy adults aged 18–59 years.^26^ However, whether it is necessary to carry out booster vaccination to strengthen immunity against SARS-CoV-2 infection and its safety and protection in young and old healthy people remains unclear.

In this study, we prepared a vaccine panel to monitor the safety and immunogenicity of three-dose booster vaccination with the Pfizer/BioNtech SARS-CoV-2 mRNA vaccine against newly emerged variant viruses, such as Delta and Omicron. In addition, we showed that booster vaccination in Japanese healthcare workers increased the breadth of humoral immunity and cross-reactivity against the SARS-COV-2 ancestral strain WK-521, as well as the newly emerged VOCs Delta and Omicron. The cytokine signatures following the three booster vaccinations were unchanged, and adverse event reports suggested that three-dose vaccinations are effective and safe.

## STAR Methods

### Study design and sample collection

This study was part of a long-term safety and immunogenicity study (LT-SI study) aimed at evaluating anti-spike antibody titers (anti-SAb) and NTs after vaccination for comparison with the adverse event rate during three-dose vaccinations with the Pfizer/BioNTech SARS-CoV-2 mRNA vaccine (Comirnaty) in healthcare workers. The vaccine was approved for emergency use with special authorization from the Japanese government in February 2021. Participants who had not been previously infected with SARS-CoV-2 were administered the SARS-CoV-2 mRNA vaccine. We collected 259 samples at each vaccination step (VAX) from the Murayama Medical Center of the National Hospital Organization (MMC-NHO). The study design is shown in **Figure 1**, and we first evaluated the anti-SAb IgG and neutralization antibody titers (**NTs**) at an average of 20 days after the first and 12 days after the second vaccination (**1st VAX** and **2nd VAX**). We prepared a post-vaccination serum panel by selecting representative samples to determine the median anti-SAb titers in each NT against WK-521 after the second vaccination (**Figure 2**). We selected 10–14 samples from the x320, x80, and x20 NTs groups and confirmed that they showed a normal distribution within each NT. The same vaccines were used after the second vaccination, including after 3 months (**2nd VAX+3M**), pre-third vaccination (**pre3rd VAX**) at 243 days after the second vaccination, and after the third vaccination (**3rd VAX**). Because of changes in the personnel within the hospital groups, we could not obtain serial data from four participants after the 2nd VAX; therefore, we added another four representatives from the x160 NT population. A total of 272 participants were enrolled in this study. The adverse events (AEs) are collected 2 weeks after the 2nd and 3rd booster vaccination using questionnaire to participants. This study was approved by the MMC-NHO (#21-01, #21-04, #21-08) and NIID Institutional Review Boards (#1270, and #1339).

**Figure 1.**
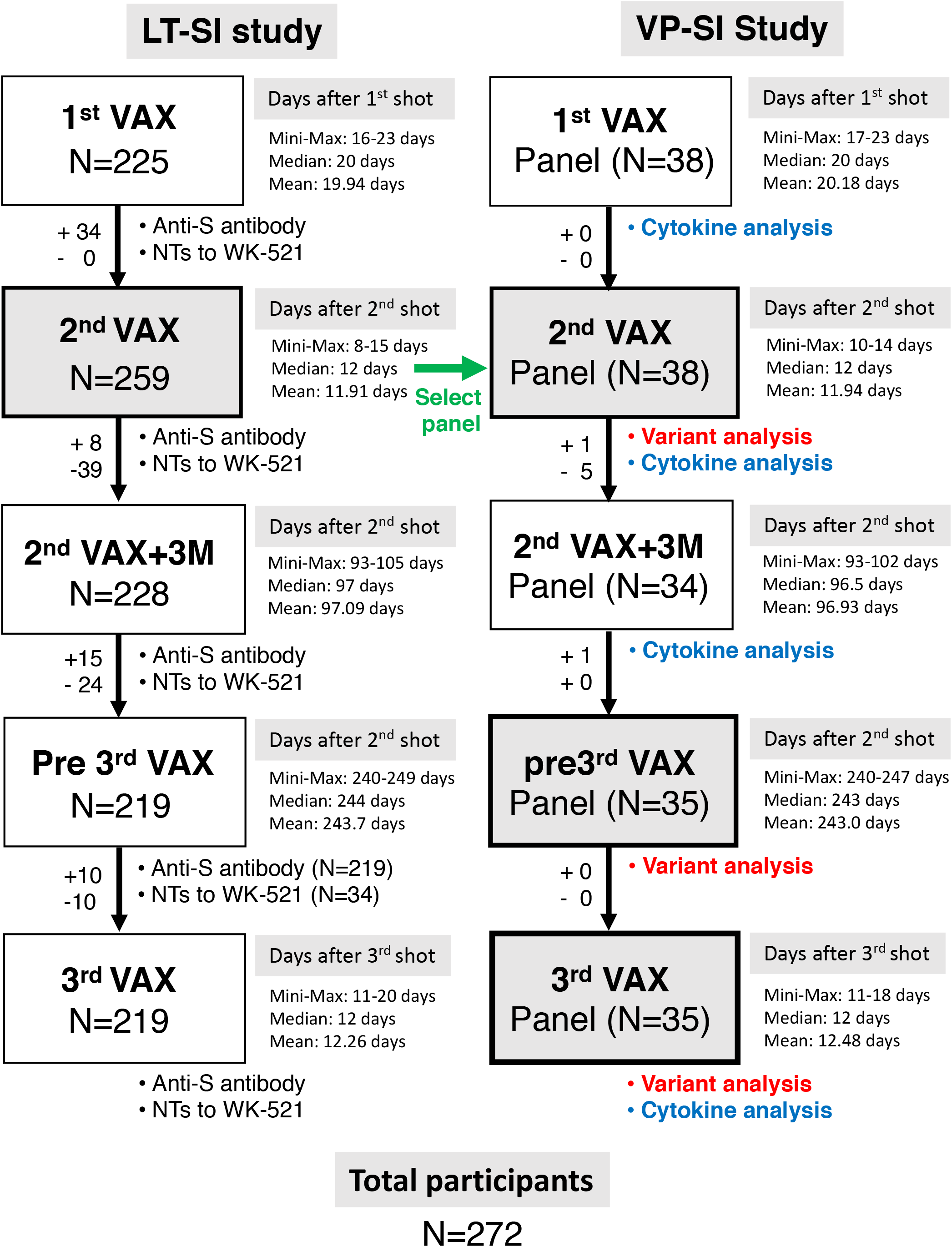
Flowchart of participant enrollment and establishment of vaccine panel. Long-term safety and immunogenicity (LT-SI) study was conducted by Murayama Medical center, National Hospital Organization in collaboration with the National Institute of Infectious Diseases. A total of 272 participants was enrolled in the LT-SI study. The vaccine panel (VP) was composed with 34–38 selected samples from participants in the LT-ST study to reflect the original population at the second vaccination.

**Figure 2.**
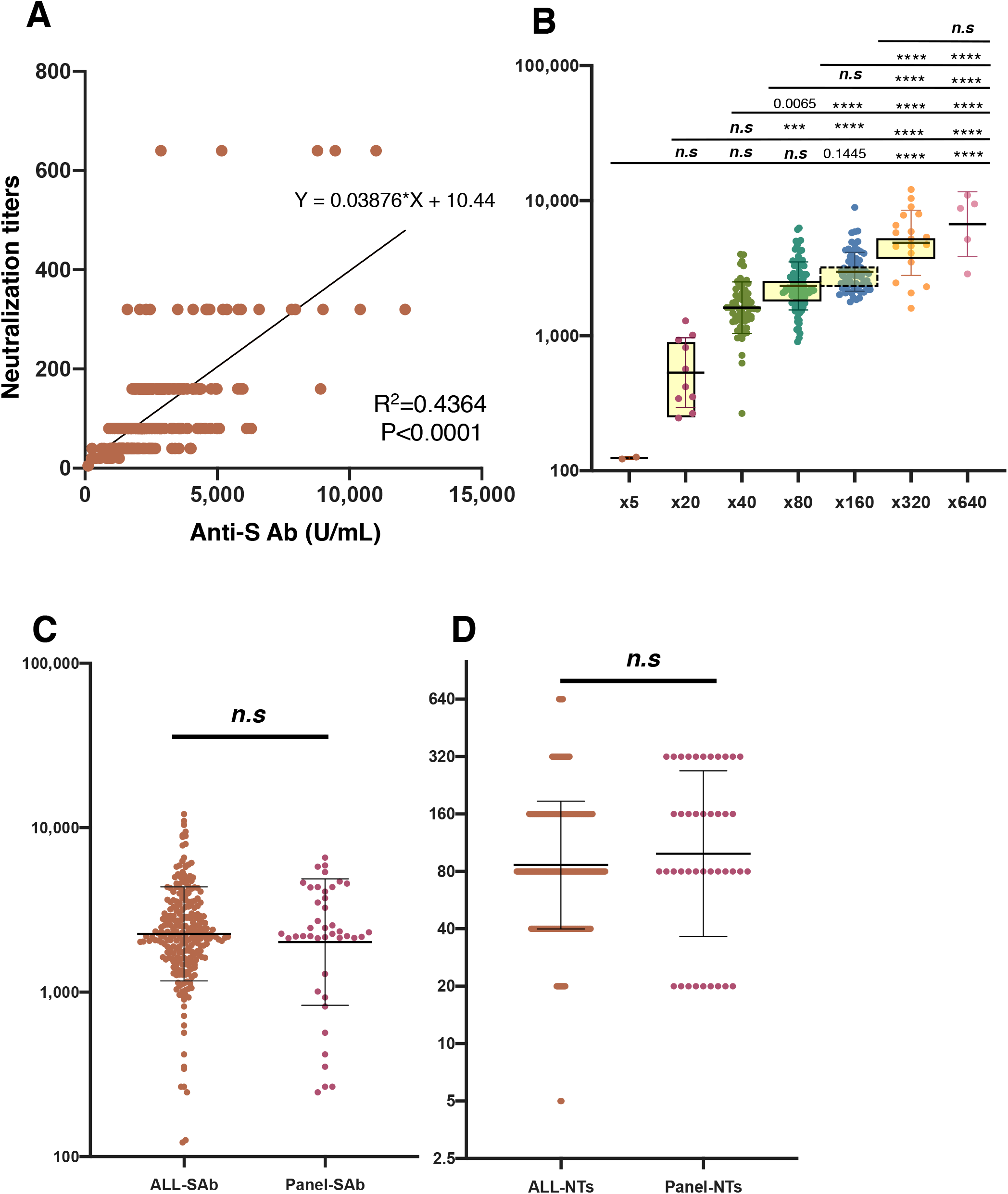
Establishment of vaccine panel. (A) Weak correlation of anti-Spike antibody (anti-SAb) and microneutralization titer (NTs) were observed at the second vaccination. (B) Anti-SAb titers were plotted in each microneutralization titer group, x5, x20, x40, x80, x160, x320, and x640, respectively. At least 10 samples showing values near mean NTs in each titer group were selected. ****<0.0001, ***<0.001, ***n*.*s***: not significant. (C) Comparison of anti-SAb in LT-SI and VP-study. (D) Comparison of NTs in LT-SI and VP-study.

### SARS-CoV-2 virus

We used the SARS-CoV-2 ancestral strain WK-521 (lineage A, GISAID ID: EPI_ISL_408667), Alpha variant QHN002 (lineage B.1.1.7, GISAID ID: EPI_ISL_804008), Gamma variant TY7-503 (lineage P.1, GISAID ID: EPI_ISL_877769), Beta variant TY8-612 (B.1.351, GISAID ID: EPI_ISL_1123289), R.1 variant TY8-524 (R.1, GISAID ID: EPI_ISL_1358213), Delta variant TY11-330 (B.1.617.1, GISAID ID: EPI_ISL_2158613), Delta variant TY11-927 (B.1.617.2, GISAID ID: EPI_ISL_2158617), Omicron variant TY38-873 (BA.1, GISAID ID: EPI_ISL_7571617), and Omicron variant TY38-871 (BA.1, GISAID ID: EPI_ISL_7571618) as live viruses. These viruses were isolated from VeroE6/TMPRSS2 cells using respiratory specimens collected from individuals screened at an airport quarantine facility in Japan at NIID with ethical approval from the Medical Research Ethics Committee of NIID for the use of human subjects (#1178). All isolated viruses were sequenced at NIID. TY38-871 was found to contain an additional R346K mutation.

### Anti-SAb evaluation

The anti-spike antibody titer was measured using the Elecsys anti-SARS-CoV-2 S assay (Roche Diagnostics International Ltd., Basel, Switzerland), which is an electrochemiluminescence immunoassay with a double-antigen sandwich design used to detect immunoglobulins in the RBD of the spike protein. This kit primarily detects IgG, IgA, and IgM. Serum samples were prepared according to the manufacturer’s instructions and analyzed using the Roche Cobas e411 platform. According to the manufacturer’s guidelines, sample values of ≥0.8□AU/mL were classified as positive for anti-SARS-CoV-2 antibodies.

### Live virus microneutralization assay

Live virus neutralization assays were performed as previously described.^27–29^ Briefly, serum samples were serially diluted (twofold dilution starting from 1:5) in high-glucose Dulbecco’s modified Eagle’s medium supplemented with 2% fetal bovine serum and 100 U/mL penicillin/streptomycin and then mixed with 100× of the median tissue culture infectious dose of SARS-CoV-2 viruses followed by incubation at 37 °C for 1 h. The virus-serum mixture was placed on VeroE6/TMPRSS2 cells (JCRB1819) seeded into 96-well plates and cultured at 37 °C with 5% CO_2_ for 5 days. After culture, the cells were fixed with 20% formalin (Fujifilm Wako Pure Chemicals, Osaka, Japan) and stained with crystal violet (Sigma-Aldrich, St. Louis, MO, USA). The mean cutoff dilution index with the >50% cytopathic effect from two multiplicate series is presented as the NT. To calculate the geometric mean of the microneutralization (MN) titers, an MN titer of 0 was considered as 2.5. All experiments using live viruses were performed in a biosafety level 3 facility at the NIID.

### Multiplex SARS-CoV-2 neutralization antibodies detection assay

SARS-CoV-2 neutralizing antibodies in serum samples were analyzed using the Bio-Plex Pro Human SARS-CoV-2 Variant Neutralization Antibody, 11-plex (Bio-Rad) according to the manufacturer’s instructions and a previous report.^30^ Briefly, 25-µL serum samples were mixed with SARS-CoV-2 antigen-coupled beads in a 96-well plate. After incubation on a shaker at 850 rpm for 30 min at room temperature, 25 µL of biotinylated detection angiotensin-converting enzyme 2 receptor was added and the mixture was incubated at 850 rpm for 30 min. After washing, 50 µL of streptavidin-phycoerythrin was added, and the mixture was incubated on a shaker at 850 rpm for 10 min at room temperature. The beads were washed and resuspended in 125 µL of assay buffer. Fluorescence intensities were measured using a Bio-Plex MAGPIX multiplex reader. Data were analyzed using Bio-Plex Manager Software version 6.0 (Bio-Rad).

### Cytokine assay

Cytokines in serum samples were analyzed using the Bio-Plex Pro Human Cytokine Screening Panel 48-plex (#12007283) (Bio-Rad) according to the manufacturer’s instructions. Briefly, 50 µL of serum samples was mixed with capture antibody-coupled beads in a 96-well plate and incubated on a shaker at 850 rpm for 1 h at room temperature. The beads were washed, mixed with 50 µL of biotinylated detection antibodies, and incubated on a shaker at 850 rpm for 30 min at room temperature. After washing, 50 µL of streptavidin-phycoerythrin reporter dye was added and incubated on a shaker at 850 rpm for 10 min at room temperature. The beads were washed and resuspended in 125 µL assay buffer. Fluorescence intensities were measured using a Bio-Plex MAGPIX multiplex reader (Bio-Rad) and the data were analyzed using Bio-Plex Manager Software version 6.0.

### Statistics

All statistical experimental designs and data analyses were performed using JMP software version 14.2.0 (SAS Institute, Cary, NC, USA) and GraphPad Prism 9.0.2 software (GraphPad, Inc., San Diego, CA, USA). Log-transformed MN antibody titers against different SARS-CoV-2 variants were compared using one-way analysis of variance with Dunn’s multiple comparisons test, Tukey’s multiple comparisons test, and paired *t*-test. Statistical significance was set to P < 0.001.

## RESULTS

### Establishment of vaccinee panel from long-term three-dose vaccination study in Japanese healthcare workers

We collected 259 samples after the 1st and 2nd VAX of Pfizer/BioNTech SARS-CoV-2 mRNA vaccine (Comirnaty) from Japanese healthcare workers (**Figure 1**). Anti-SAb titers were measured using the Roche Elecsys ® Anti-SARS-CoV-2 (RUO), and NTs were measured using live virus for the original ancestral vaccine strain, WK-521, of SARS-CoV-2, as described previously.^27^ Although anti-SAb and NTs showed a weak correlation at the second vaccination in our preliminary analysis (**Figure 2A**), each anti-SAb titer was normally distributed in the NT, x20, x40, x80, x160, x320, and x640 groups. We selected 10–14 representative samples from around the mean value of anti-SAb for each NT (**Figure 2B**). The participants in the LT-SI study are listed in **Table 1**, and the characteristics of the selected vaccine panel study (**VP-SI study**) are described in **Table 2**. We selected 34–38 samples as reflected in the original LT-SI study. We also included patients who developed adverse events, including fever, headache, fatigue, and injection site pain, which were similar to those in the LT-SI study at 12 days after the second (2nd VAX) and third vaccinations (3rd VAX). There was no significant difference in the anti-SAb and NT distributions between the LT-SI and VP-SI groups (**Figure 2C** and **D**).

**Table. 1.** Characteristics of study subjects vaccinated with Pfizer/BioNTech BNT162b2 SARS-CoV-2 mRNA.

|  | Male |  | Female |  | Total |  |
| --- | --- | --- | --- | --- | --- | --- |
| Total number | 103 | 38% | 169 | 62% | 272 |  |
| Age (years), mean | 42.1 |  | 40 |  | 40.8 |  |
| Age group (years) |  |  |  |  |  |  |
| 20-29 | 15 | 25% | 45 | 75% | 60 | 22% |
| 30-39 | 33 | 48% | 36 | 52% | 69 | 25% |
| 40-49 | 26 | 34% | 51 | 66% | 77 | 28% |
| 50-59 | 21 | 43% | 28 | 57% | 49 | 18% |
| 60-64 | 8 | 47% | 9 | 53% | 17 | 6% |

**Table. 2.** Characteristics of selected vaccinees panel with Pfizer/BioNTech BNT162b2 SARS-CoV-2 mRNA.

|  | Male |  |  |  | Female |  |  |  | Total |  |  |  |
| --- | --- | --- | --- | --- | --- | --- | --- | --- | --- | --- | --- | --- |
|  | 2ndVAX |  | 3rdVAX |  | 2ndVAX |  | 3rdVAX |  | 2ndVAX |  | 3rdVAX |  |
|  | N | % Total | N | % Total | N | % Total | N | % Total | N | % Total | N | % Total |
| Total | 17 | 45% | 16 | 46% | 21 | 55% | 19 | 54% | 38 |  | 35 |  |
| Age group (years) |  |  |  |  |  |  |  |  |  |  |  |  |
| 20-29 | 3 | 33% | 3 | 33% | 6 | 67% | 6 | 67% | 9 | 24% | 9 | 26% |
| 30-39 | 7 | 64% | 7 | 70% | 4 | 36% | 3 | 30% | 11 | 29% | 10 | 29% |
| 40-49 | 0 | 0% | 0 | 0% | 6 | 100% | 6 | 100% | 6 | 16% | 6 | 17% |
| 50-59 | 5 | 56% | 4 | 57% | 4 | 44% | 3 | 43% | 9 | 24% | 7 | 20% |
| 60-64 | 2 | 67% | 2 | 67% | 1 | 33% | 1 | 33% | 3 | 8% | 3 | 9% |
| Adverse event |  |  |  |  |  |  |  |  |  |  |  |  |
| Fever |  |  |  |  |  |  |  |  |  |  |  |  |
| - | 5 | 38% | 5 | 38% | 8 | 62% | 8 | 62% | 13 | 34% | 13 | 37% |
| + | 12 | 48% | 11 | 50% | 13 | 52% | 11 | 50% | 25 | 66% | 22 | 63% |
| 37°C | 5 | 45% | 8 | 67% | 6 | 55% | 4 | 33% | 11 | 29% | 12 | 34% |
| 38°C | 5 | 56% | 3 | 33% | 4 | 44% | 6 | 67% | 9 | 24% | 9 | 26% |
| 39°C | 2 | 40% | 0 | 0% | 3 | 60% | 1 | 100% | 5 | 13% | 1 | 3% |
| Headache |  |  |  |  |  |  |  |  |  |  |  |  |
| - | 10 | 63% | 9 | 69% | 6 | 38% | 4 | 31% | 16 | 42% | 13 | 37% |
| + | 7 | 32% | 7 | 32% | 15 | 68% | 15 | 68% | 22 | 58% | 22 | 63% |
| Fatigue |  |  |  |  |  |  |  |  |  |  |  |  |
| - | 6 | 60% | 7 | 70% | 4 | 40% | 3 | 30% | 10 | 26% | 10 | 29% |
| + | 11 | 39% | 9 | 36% | 17 | 61% | 16 | 64% | 28 | 74% | 25 | 71% |
| Injection site pain |  |  |  |  |  |  |  |  |  |  |  |  |
| - | 4 | 50% | 0 | NA | 4 | 50% | 0 | NA | 8 | 21% | 0 | NA |
| + | 13 | 43% | 16 | 46% | 17 | 57% | 19 | 54% | 30 | 79% | 35 | 100% |

### Three-dose vaccinations induce higher anti-SARS-CoV-2 spike IgG and neutralization antibodies

In our VP-SI study, the anti-SAb titers were significantly increased (x46) after the second and third Pfizer/BioNTech SARS-CoV-2 mRNA vaccine (**Figure 3A**) compared to those after the first vaccine. Although anti-SAb levels slightly decreased at 97 (2nd VAX+3M) and 243 days (pre3rd VAX) after the second vaccination, anti-SAb levels remained significantly higher than those after the 1st VAX. A similar trend was observed in the NT group (**Figure 3B**). The NTs significantly increased (x19) after the 2nd VAX. NTs also decreased with the 2nd VAX+3M and pre3rd VAX. Although NTs at the pre3rd VAX were not higher than those at the 1st VAX, NTs at the 2nd VAX+3M were higher than those after the 1st VAX. The third dose of the mRNA vaccine dramatically and significantly increased both anti-SAb (x62) and NTs (x44) compared to that of the pre3rd VAX.

**Figure 3.**
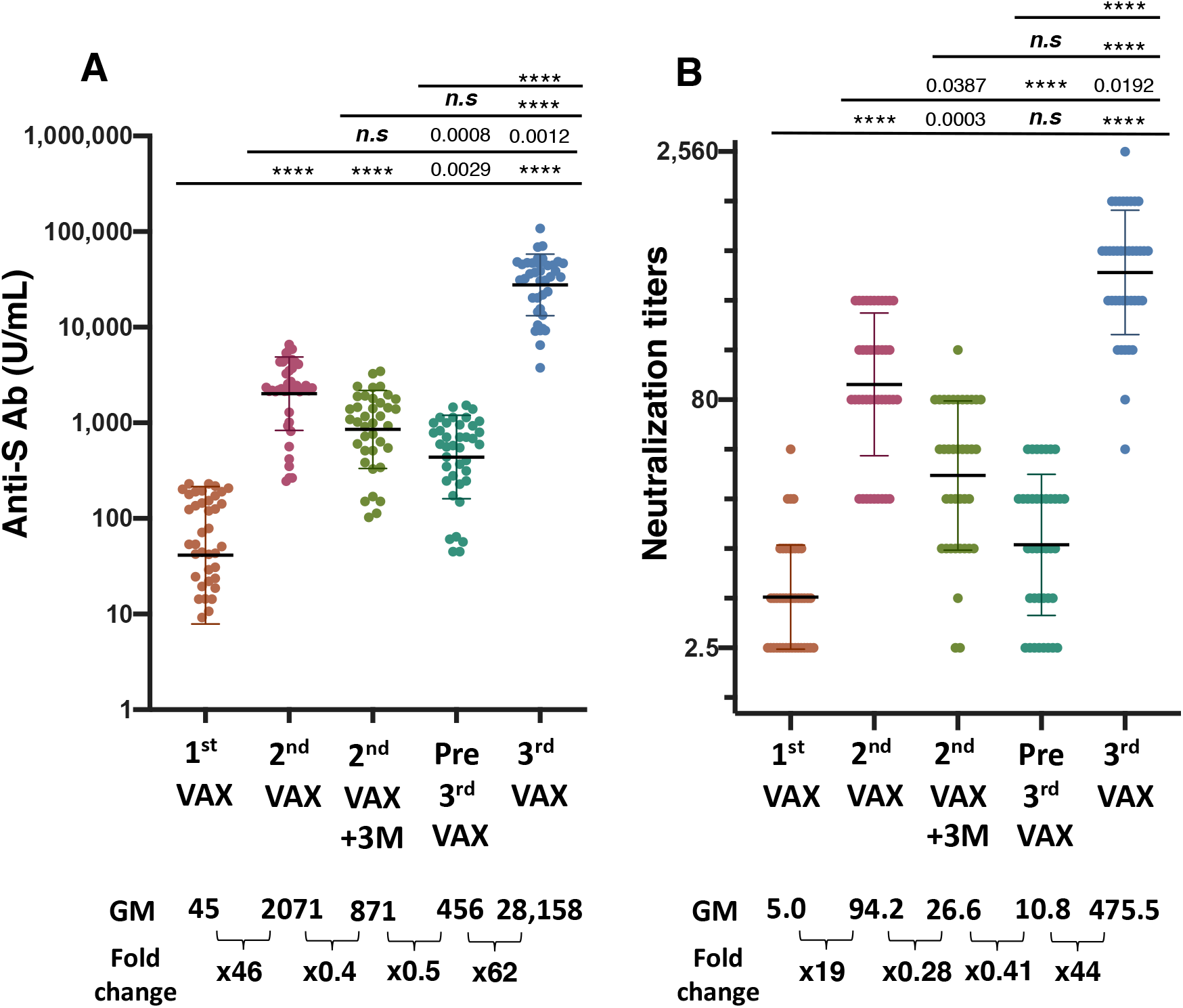
Anti-spike antibody titer (anti-SAb) and microneutralization titer (NTs) after three-dose vaccination with Pfizer/BioNTech mRNA vaccine. Antibody responses at 12 days after the first vaccination (1stVAX), 20 days after the second vaccination (2nd VAX), 3 months after the second vaccination (2ndVAX+3M), 244 days after the second vaccination (pre3rd VAX), and 12 days after the third vaccination (3rd VAX) were plotted. (A) Anti-SAb (U/mL) was measured using a Roche Elecsys anti-SARS-CoV-2 S assay. (B) Microneutralization titers were measured with live virus, WK-521, and the dot plot shows the data of each participant. Geometric mean and SD are indicated. Fold-change is the value compared to the previous timepoint. ****<0.0001, ***<0.001, ***n*.*s***: not significant.

### Second dose of vaccination was effective for some variants but not for Omicron

The Pfizer mRNA vaccine shows 95% effectiveness against the ancestral strain, WK-521.^12^ However, some groups reported that NTs for the Beta and Kappa strains were decreased^35–37^ after two-dose Pfizer mRNA vaccination, as well as after other approved vaccines such as the Moderna mRNA^38^ and AstraZeneca adenovirus vector DNA vaccines.^39^ Thus, we tested whether two-dose vaccinations exhibited neutralization activity against some SARS-CoV-2 VOCs, including the recently emerged Omicron strain (**Figure 4A**). Two doses of the Pfizer mRNA vaccine significantly decreased the NTs for the Beta and Kappa strains, as well as the Delta variant, which was endemic in Japan as of December 2021. Two Omicron strains, BA.1, categorized as TY38-873 and TY-871, showed dramatically decreased NTs after two doses of Pfizer mRNA vaccination.

**Figure 4.**
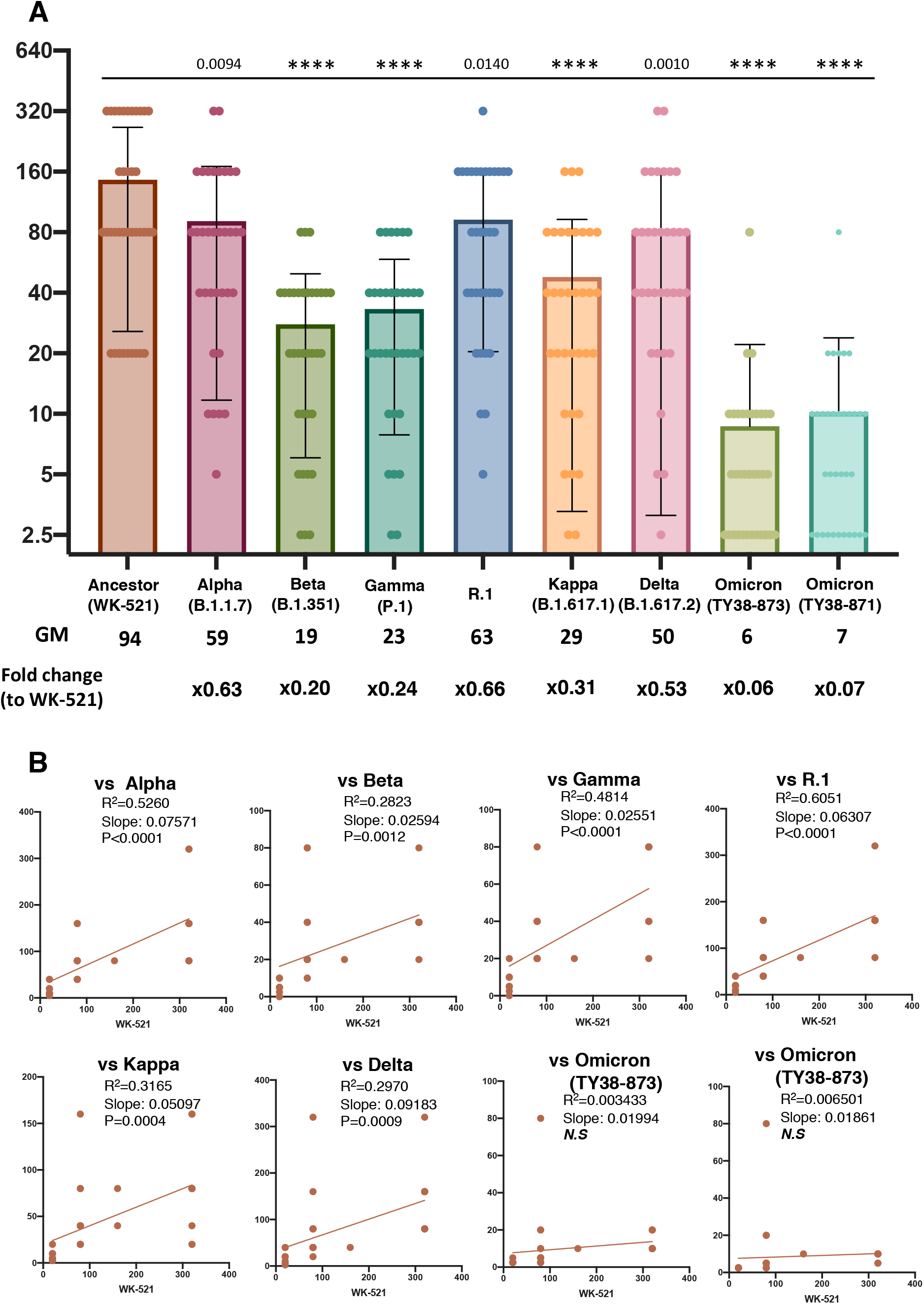
Comparison of neutralization ability in vaccinee sera from two-dose vaccination. (A) Microneutralization titer of 2nd VAX in WK-521, Alpha, Beta, Gamma, R.1, Kappa, Delta, two Omicron variant are shown. ***<0.0001, ***<0.001, ***n*.*s***: not significant. (B) **Cross-reactivity of NTs in WK-521 and SARS-CoV-2 variant**. Linear regression analysis of wild type versus each variant. NTs in Wild type Wk-521 correlated with Alpha, Beta, Gamma, R.1, Kappa, Delta. No correlation to Omicron variant.

To characterize the neutralization patterns observed between individuals who were vaccinated with the second dose of each Pfizer mRNA vaccine, we directly compared the wild-type NTs of these two groups of samples against VOCs (**Figure 4B**). We found that wild-type NTs from individuals administered their second vaccination dose weakly correlated with Alpha, Beta, Gamma, R.1, Kappa, and Delta and did not correlate with two Omicron variant cross-neutralization at the 2nd VAX. These data suggest that second-dose vaccination induced a high NT against the vaccine strain WK-521 but not against the Omicron variant.

### Cytokine signature was stable during vaccinations 1–3

The Pfizer/BioNtech mRNA vaccine is a new modality used in Japan and other countries. Although this vaccine has been shown to be effective, there are some safety concerns by the healthy general population given its emergency authorization. In this LT-SI study, we collected information on adverse events and summarized vaccine safety data at each time point after vaccination (**Table 2**). Briefly, there were no serious adverse events associated with this vaccine compared to those in previous phase II/III clinical studies in Japan and other countries. In addition, we collected 2nd VAX, 2nd VAX+3M, and 3rd VAX samples to measure the 48 cytokine profiles. During vaccination, enhanced cytokine production, including some inflammatory cytokines, was not observed at any time point (**Figure 5**). The cytokine levels of eotaxin in the 3rd VAX group were significantly higher than those in the 2nd VAX and 2nd VAX+3M groups. The cytokine levels of CTACK, cutaneous T cell-attracting chemokine in the 2nd VAX+3M group were significantly lower than those in the 1st and 2nd VAX groups. The cytokine levels of macrophage inhibitory factor and macrophage inhibitory protein-1β in the 3rd VAX group were significantly lower than those in the 1st VAX group.

**Figure 5.**
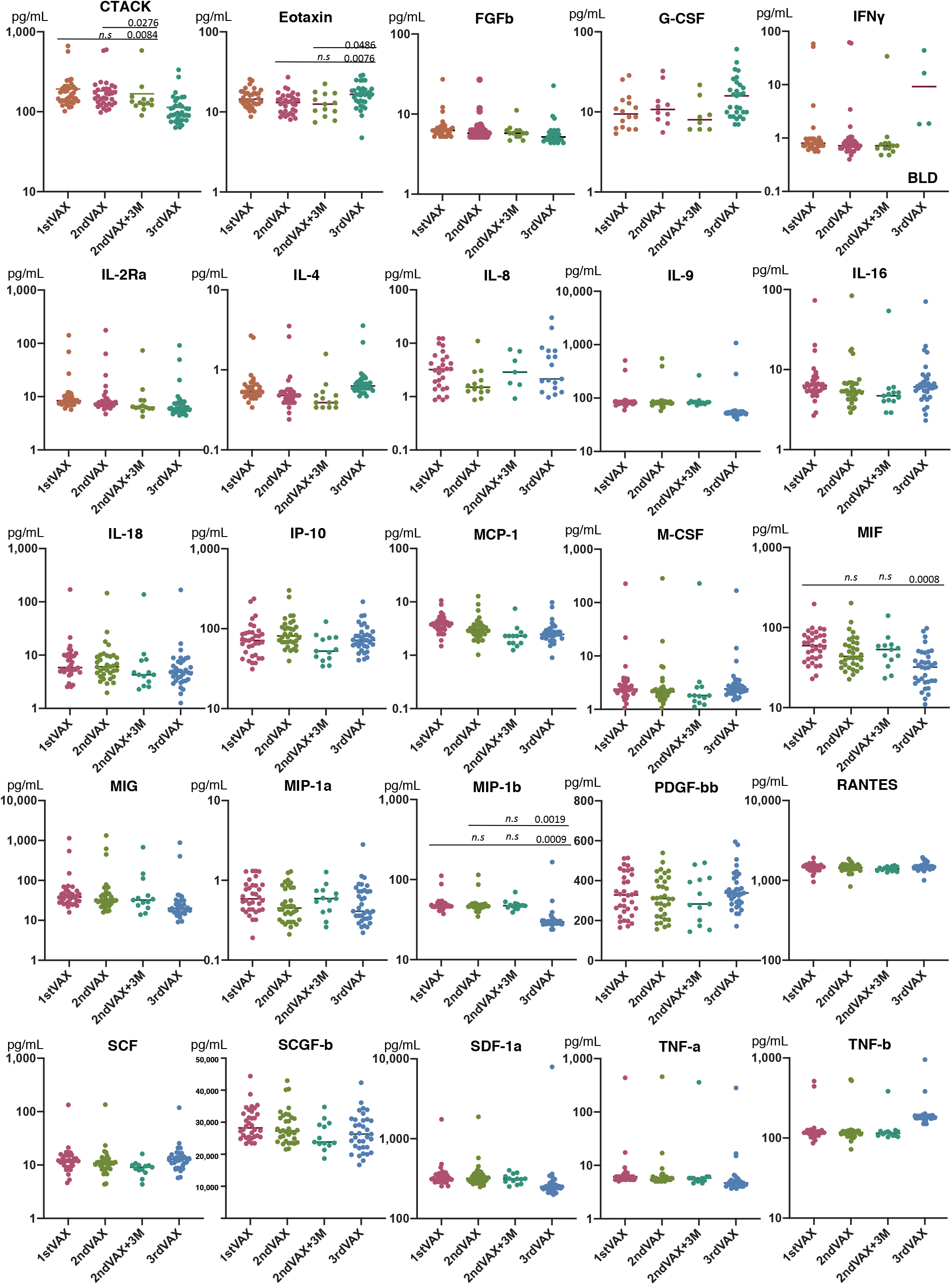
Cytokine signature during three-dose vaccination with Pfizer/BioNTech mRNA vaccine. Cytokine production at 3rd VAX was increased in cutaneous T cell-attracting chemokine (CTACK), eotaxin, macrophage inhibitory factor (MIF), and macrophage inhibitory protein (MIP)-1β, ***<0.0001, ***<0.001, ***n*.*s***: not significant or no description on the data.

When we divided the subjects into adverse event-positive and adverse event-negative groups and high fever and low fever groups, we observed no differences in the cytokine signatures between the different groups (**data not shown**). There was no correlation between body temperature and cytokine levels in the 2nd and 3rd VAX groups (**data not shown**). In addition, we analyzed the correlation between anti-SAb and each cytokine levels and found that only the eotaxin level was weakly correlated with the anti-SAb titer in the 3rd VAX group.

### Three doses of vaccination increase both anti-Sab and NTs that sufficiently cross-react with some variants

In Japan, the third booster vaccination was started in November 2021, only for healthcare workers. We collected pre3rd VAX and 3rd VAX samples from the same participants in our panel. We first compared NTs with those of the original vaccine strain, WK-521. NTs in WK-521 cells were significantly higher than those in the 2nd VAX samples (**Figure 6A**). Some values exceeded x2,560, and the geometric mean of the NTs was x476 at the 3rd VAX. Subjects injected with three vaccination doses showed a 43.95-fold higher NT compared to those in the pre3rd VAX group. A similar result was obtained for the Delta variant (**Figure 6B**). NTs were x238 in the 3rd VAX group, showing a 36.04-fold increase compared to in the pre3rd VAX group. In the most recently identified VOC strain, Omicron and NTs in the 2nd VAX were x6 and NTs in the 2nd VAX+3M were x3, which were lower than those of the WK-521 and Delta variants (**Figure 6C**). Most NTs in the pre3rd VAX were below the detection limit (x2.5). However, NTs in the 3rd VAX were increased by 33.29-fold at the 3rd VAX, which was higher than the NTs in WK-521 at the 2nd VAX. A similar result was obtained using a commercially available multiplex SARS-CoV-2 neutralization antibody detection assay kit. Neutralizing antibodies against wild-type S1, Alpha S1, BetaS1, and D614G S1 were significantly increased by 1.96–2.98-fold compared to in the 2nd VAX group. Neutralizing antibodies against the wild-type RBD, Gamma RBD, Kappa RBD, Epsilon RBD, N501Y RBD, K417RBD, and E484K RBD significantly increased by 2.35–3.15-fold compared to those after the 2nd VAX (**Figure 6D**).

**Figure 6.**
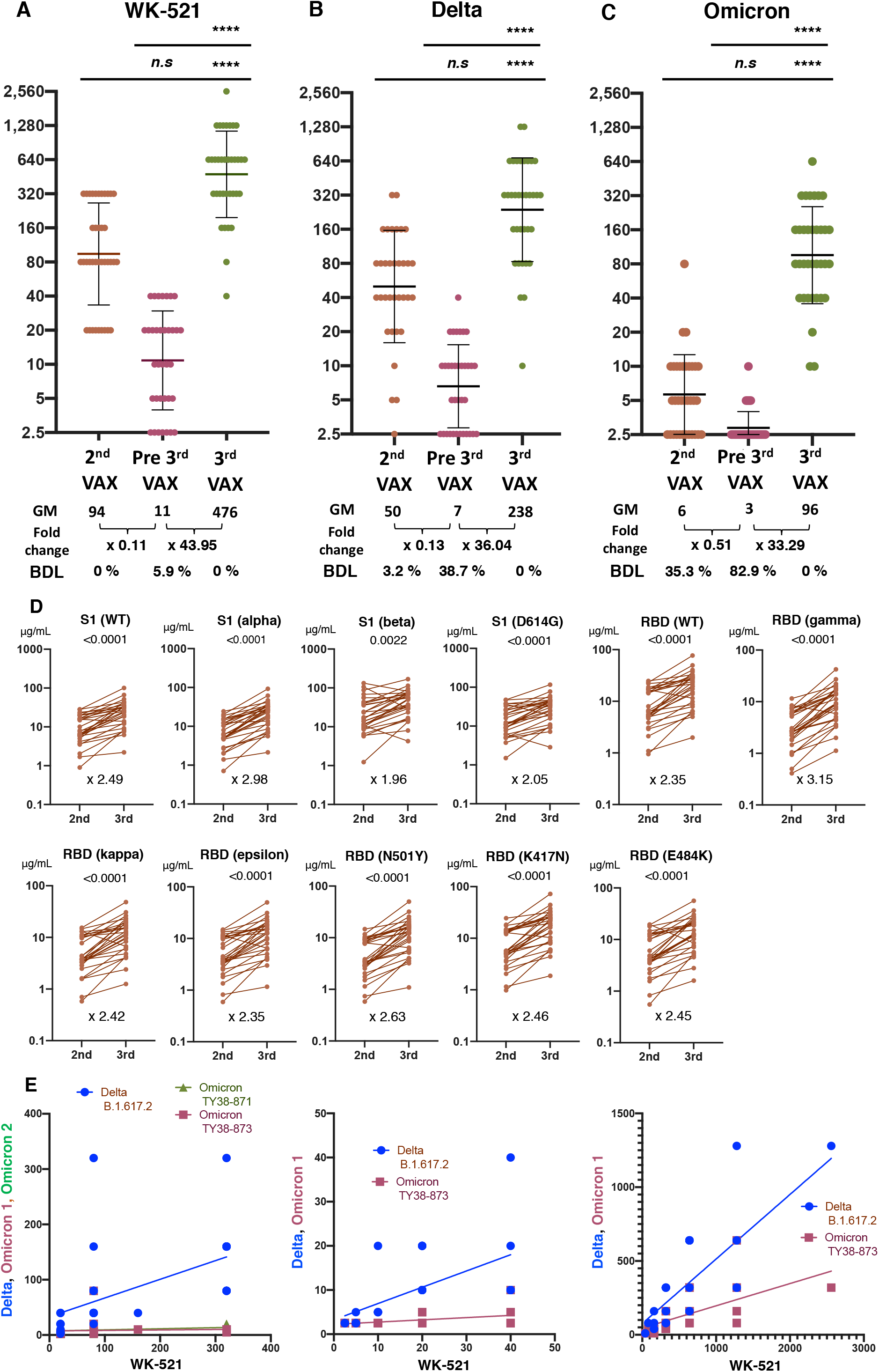
Neutralization antibody against WK-521, Delta variant, and Omicron variant after three-dose vaccination. **(A)** Neutralization titers (NTs) against WK-521. **(B)** NTs against Delta variant. **(C)** NTs against Delta variant. (**D**) Comparison of neutralization antibody measured with the Bio-Plex Pro Human Cytokine Screening Panel, 48-plex. **(E)** Linear regression analysis of wild-type versus Omicron at 2nd VAX and pre3rd VAX and 3rd VAX. NTs in wild-type WK-521 correlated with Omicron at booster vaccination.

### Three-dose booster vaccination increased breadth of humoral immunity and cross-reactivity against SARS-CoV-2 variant

To characterize the neutralization patterns between individuals who were booster-vaccinated with each Pfizer mRNA vaccine, we directly compared the wild-type NTs of these two groups of samples against the Delta and Omicron strains (**Figure 6E**). We found that wild-type NTs from individuals administered their second dose vaccination were weakly correlated with Delta variant cross-neutralization and not correlated with TY38-871 Omicron variant cross-neutralization or TY38-873 Omicron variant cross-neutralization in the 2nd VAX and pre3rd VAX groups (data not shown), respectively. In contrast, wild-type neutralization of boosted individuals was correlated with Delta and Omicron variant cross-neutralization. These data suggest that booster vaccination induces not only higher NTs against the vaccine strain WK-521 but also the breadth of humoral immunity and cross-reactivity against the highly mutated SARS-CoV-2 Omicron variant.

## DISCUSSION

We first determined the changes in anti-SAb and NTs against Omicron variants in Japanese healthcare workers during three-dose vaccination with the Pfizer/BioNTech SARS-CoV-2 mRNA vaccine (Comirnaty). The second vaccine dose dramatically increased both the anti-SAb and anti-NT against the vaccine strain WK-521. The effectiveness of this vaccine has already been established,^12^ preventing more than 95% of COVID-19 cases in phase II/II clinical studies. Since SARS-CoV-2 WK-521 was first isolated, many additional variants have been identified and evaluated by the WHO to determine their virological characteristics, pathogenicity, and impact on society; these variants have been designated as VOCs, variants of interest, and variants under monitoring (WHO website). Since 2020, the WHO has named Alpha, Beta, Gamma, Delta, and Omicron as VOCs. The Kappa variant used in this study was first categorized as a VOC, after which the WHO excluded the Kappa variant as a VOC. The R.1 variant used in this study was first identified as Pangolin B.1.1.316 containing an E484K mutation in the spike region, enabling escape vaccine-induced immunity, in February 2021 in Japan. R.1 variants were also first categorized as VOCs and then categorized as variants of interest. In this study, we compared NTs with VOCs after the second dose of vaccination. Our results agree with those of a previous study showing that NTs were severely reduced for Beta and Gamma strains, which exhibit three-vaccine escape mutations in the spike RBD domain.^19^ Similarly, NTs against Delta and Gamma were slightly decreased compared to those against the ancestral strain, WK-521. In November 2021, the SARS-CoV-2 Omicron variant (BA.1/B.1.1.529), which contains more than 15 mutations in the spike RBD and overlap the three sites mutated in Gamma and Beta (K417, E484, and N501), including vaccine escape mutations, emerged in Botswana. This variant was reported to the WHO in November 2021, as a novel VOC, harboring up to 36 mutations with immune-evasive potential in the spike protein, the target of neutralizing antibodies, and spread rapidly worldwide. These mutations in Omicron indicate the potential for antibody escape from mRNA vaccine-induced immunity and are more resistant to neutralization by the mRNA vaccine sera.^31^ In Japan, the first Omicron case was reported in December 2021. We isolated and sequenced two Omicron strains with R346K mutations in the spike RBD domain, which have been observed at relatively low frequency (<10%) within the Omicron lineage, but which are known as key targets for escaping neutralizing monoclonal antibodies such as AZD1061/cilgavimab.^32–34^ Using the two newly isolated Omicron strains, we observed a severe reduction in NTs for the Omicron strain. The serum of most participants (35%) was detected at the detection limit (x2.5), indicating low or no cross-reactivity against Omicron even at 10–12 days after the second vaccination. Both NTs in the WK-521, Delta, and Omicron stains drastically decreased at 244 days after the second vaccination, as indicated in **Figures 3 and 6**. However, the percentage of NTs around the detection limit (x2.5) was 6% in the WK-521 strain, 39% in the Delta strain, and 83% in the Omicron strain. These NT monitoring data clearly show that the effectiveness of the second vaccine dose was not ensured at 243 days or more. Cellular immunity and natural immunity are involved in protecting against SARS-CoV-2 infection after mRNA vaccination and^43, 44^ disease progression; thus, the NT data in the serum may not fully reflect actual immunity against SARS-CoV-2. Therefore, many countries recommend booster vaccination 6 months after the second vaccination. In our study, three-dose vaccination significantly and dramatically increased anti-SAb and NTs for WK-521, Delta, and Omicron. NTs for Delta in the third booster vaccination sample were higher than those for WK-521 in the 2nd VAX. NTs for Omicron in the third boosted vaccination sample were the same as those in WK-521 at the 2nd VAX. These data suggest that similar and >95% effectiveness against WK-521 can be expected with three-dose vaccinations for both the Delta and Omicron variants. In addition, third booster vaccination induced humoral immunity and cross-reactivity to the Delta and Omicron variants. Several studies have suggested that mRNA vaccination induces a humoral immune response and immune memory.^43, 44^ Thus, booster vaccination may reduce the emergence of new variants.

In addition to immunogenicity, safety is a critical issue, as some people consider that emergency use authorization means that complete safety research has not been performed, resulting in vaccine hesitancy. A phase II/III study suggested that the booster shot does not change the adverse event rate or specific adverse events in short-term analysis.^45^ During three-dose vaccination, abnormal cytokine production was not observed in healthy participants or in those who developed fever, headache, fatigue, or injection site pain. Only eotaxin levels increased at the 3rd VAX compared to at the 2nd VAX. Eotaxin, a chemokine ligand for CCR3, is a chemoattractant for eosinophils, basophils, and Th2 lymphocytes and is released from endothelial and epithelial cells. Eotaxin recruits eosinophils to the site of inflammation and releases reactive oxygen species, causing tissue damage during chronic inflammatory responses. Several studies have shown that eotaxin levels increased after the smallpox vaccination according to the adverse event.^46^ Thus, eotaxin levels may be related to adverse events such as injection site pain and redness after the 3rd VAX. However, damage-associated inflammatory cytokine levels induced by eosinophils were unchanged in our samples, and key pathways involving eotaxin, such as interferon-gamma and interleukin-4 were unchanged. These data suggest that eotaxin alone is not an indicator of serious adverse events. Multiple pathway analyses are needed to determine the relationship between cytokine levels and adverse events. Overall, the cytokine data suggest no signs of adverse events.

Taken together, booster vaccination induced not only an increase in the anti-Spike IgG titer, but also an increase in the breadth of humoral immunity and cross-reactivity against newly emerged Omicron variants without causing specific adverse events or abnormal cytokine production.

## Data Availability

All data produced in the present study are available upon reasonable request to the authors.

## ACKNOWLEDGEMENTS

We thank Dr. **Noriyo Nagata, Shinji Watanabe, and Seiichiro Fujisaki** of the National Institute of Infectious Diseases (NIID) for their technical support with the micro-neutralization assay using live viruses. We also thank the healthcare workers who participated in this study at the MMC-NHO. **Keiko Imai** and **Mieko Ishi** of the NIID provided technical support for this study. This study was funded by AMED under grant number JP21fk0108104 and JP21mk0102146. The content is solely the responsibility of the authors and does not necessarily reflect the official views of the NIID and Ministry of Health Japan Authority.

## AUTHOR CONTRIBUTIONS

YS, KN, TM, YY, and IH conceptualized the study; YS, KN, and TM designed the study; YS, KN, HM and TM performed experiment and analyzed the data; SF and KM provided isolated SARS-CoV-2 variants and consulted on the analysis and results; TM wrote the manuscript, AW, NN, KS,TK and YY curated the clinical data and reviewed results, SM, YT, and TS provided monoclonal antibody and rabbit polyclonal antibody against SARS-CoV-2 RBD and consulted on the experiment and analysis; all authors reviewed the manuscript.

## ETHICS STATEMENT

Deidentified and residual specimens, which were tested for anti-spike antibody at LSI Medience, were used in this study. This study was approved by the MMC-NHO (#21-01, #21-04, #21-08) and NIID Institutional Review Boards (#1270 and #1339) in accordance with the Helsinki Declaration.

